# Prognostic factors of clinical responses in patients with advanced-stage intrahepatic cholangiocarcinoma following *Atractylodes lancea* administration: A phase 2A clinical trial

**DOI:** 10.1101/2023.03.28.23287855

**Authors:** Teerachat Saeheng, Juntra Karbwang, Anurak Cheomung, Nisit Tongsiri, Tullayakorn Plengsuriyakarn, Kesara Na-Bangchang

## Abstract

A statistical model is essential in determining the appropriate predictive indicators for therapies in many types of cancers. Predictors have been compared favorably to the traditional systems for many cancers. Thus, this study has been proposed as an alternative or a new standard approach. A recent study on the clinical efficacy of *Atractylodes lancea* (Thunb) DC. (AL) revealed the higher clinical benefits in patients with advanced-stage intrahepatic cholangiocarcinoma (ICC) treated with AL compared with standard supportive care. we investigated the relationships between clinical efficacy and pharmacokinetic parameters of serum bioactivity of AL and its active constituent “atractylodin” and determined therapeutic ranges. Cox proportion hazard model and Receive Operating Characteristic (ROC) were applied to determine the cut-off values of AUC_0-inf_, C_max_, and C_avg_ associated with therapeutic outcomes. Number-need to be treated (NNT) and relative risk (RR) was also applied to determine potential predictors. The AUC_0-inf_ of total AL bioactivity of> 96.71 µg*h/ml was identified as a promising predictor of disease prognosis, *i*.*e*., progression-free survival (PFS) and disease control rate (DCR). C_max_ of total AL bioactivity of>21.42 was identified as a predictor of the prognosis of death. The therapeutic range of total AL bioactivity for PFS and DCR is 14.48-65.8 µg/ml, and for overall survival is 10.97-65.8 µg/ml. The predictors of ICC disease prognosis were established based on the pharmacokinetics of total AL bioactivity. The information could be exploited to improve the clinical efficacy of AL in patients with advanced-stage ICC. These predictors will be validated in a phase 2B clinical study.

## 1. Introduction

Intrahepatic cholangiocarcinoma (ICC), a biliary tract cancer, accounts for 20% of primary liver cancers [1]. It is recognized as a deadly gastrointestinal cancer with a poor disease prognosis due to the lack of effective biomarkers for early diagnosis and effective treatment. With advanced disease, patients receiving first-line chemotherapy have a 5-year overall survival (OS) rate of lower than 20% [1]. High rate resistance of ICC to first-line chemotherapy results in disease progression among the treated patients [2]. Research and development of effective alternative therapies are needed to tackle this fatal disease and improve the quality of life in patients with advanced-stage ICC.

*Atractylodes lancea* Thunb. (DC.) (AL), and the two active constituents --atractylodin (ATD) and β-eudesmol, have been shown to be promising candidates for CCA in a series of non-clinical and clinical studies with acceptable toxicological profiles [3]. Recently, a phase-I randomized controlled trial study of AL has demonstrated immunomodulatory effect of AL, *i*.*e*., the suppression of the production of inflammatory cytokines, and the promotion of peripheral immunostimulant cells [4]. Based on the classification of the immune subtypes (distinct composition and functions of tumor microenvironment (TME), AL would support the new paradigm for the treatment of ICC in promoting the recruitment of immune cells in TME [5-10]. A phase 2A study was further conducted to assess the clinical efficacy and safety of AL in patients with advanced-stage ICC [11]. The present study aimed to 1) determine the relationship between total AL bioactivity or ATD and clinical responses (*i*.*e*., disease control rate, tumor size progression, progression-free survival, and overall survival) following treatment with capsule formulation of the standardized AL extract (CMC-AL) in patients with advanced-stage ICC, and 2) determine therapeutic ranges of AL.

## 2. Materials and methods

### 2.1 Study design

The study is a part of a single center, open-label, randomized controlled, phase 2A trial which was conducted at Sakhon Na-Kon Hospital, Sakhon Na-Kon province, Thailand (n=48) [11]. The clinical trial is registered at the Thai Clinical Trials Registry (No. TCTR20210129007). The study was approved by the Ethics Committee of Thammasat University (No. 003/2564) and Sakhon Na-Kon Hospital (No. 049/2563). The study was conducted in accordance with Good Clinical Practice (GCP) guidelines and the Declaration of Helsinki [11].

Patients with advanced-stage ICC were randomized to three groups. Groups 1 (n=16) and 2 (n=16) to receive CMC-AL with different dosage regimens in conjunction with standard supportive care, while group 3 patients (n=16) received only standard supportive care. Block randomization was applied to patients for enrollment in each group. The total study period was 4 months. It is noted that this study is a phase 2A clinical trial, the lowest number of participants for a pharmacokinetic analysis for a model prediction is at least 12 peoples for the prove of concept.

### 2.2 Patients

Eligible patients were aged ≥ 18 years with advanced-stage (unresectable ICC or metastatic ICC), with Eastern Cooperative Oncology Group (ECOG) Performance Status of 0-2, serum carcinoembryonic antigen (CEA) of > 5.2 ng/ml, tumor lesion size of > 20 mm (at least one lesion), no history of treatment with chemotherapy or radiotherapy, normal cardiac function and electrocardiogram (ECGs), adequate bone marrow functions (polymorphonuclear cells of ≥ 1,500 cells/mm^3^, platelets of ≥ 100,000 cells/mm^3^, hemoglobin of ≥ 8.0 g/dl, and normal blood coagulation or bleeding), normal liver functions (bilirubin of < 1.5 of the upper limit of normal (ULN), alkaline phosphatase, alanine aminotransferase, or aspartate aminotransferase of < 5 of ULN), and effective communication ability. Exclusion criteria were pregnancy or lactation, hypersensitivity or idiosyncratic reaction to herbal products or medicines, current or previous diagnosis of other cancers within five years, gastrointestinal abnormality, immune deficiency, and participation in other studies before study in the last three months. All patients provided informed consent before enrollment.

### 2.3 Treatment

Group 1: daily dose of 1,000 mg CMC-AL (9 capsules) for 90 days, in conjunction with standard supportive care (n=16).

Group 2: daily dose of 1,000 mg CMC-AL (9 capsules) for 14 days, followed by 1,500 mg (14 capsules) for 14 days, and 2,000 mg (18 capsules) for 62 days, in conjunction with standard supportive care (n=16).

Group 3: standard supportive care alone (n=16).

Each capsule contained 2.45 mg and 4.06 mg of atractylodin and β-eudesmol, respectively. The drug was given in the morning, two hours before meal.

Pharmacokinetic study

The pharmacokinetic study was conducted in 32 patients. Blood samples (0, 0.25, 0.5, 1, 1.5, 2, 2.5, 3, 4, 5, 6, 8 hours, 5 ml each) were drawn from the patients on day 1 for group 1, and day 14 and day 28 for group 2.

### 2.4 Bioanalysis of atractylodin and serum total bioactivity of A. Lancea

HPLC-UV for determination of atractylodin

Atractylodin concentrations in plasma samples were determined using high performance liquid chromatography (HPLC) according to the previously reported method [12].

Bioassay for determination of total bioactivity of A. lancea

The bioassay method for determination of serum total AL bioactivity was according to the method of Choemung et al., 2022 [13].

### 2.5 Outcomes

The area under the curve (AUC_0-t_) (from zero to time t), AUC_0-inf_ (from zero to infinity), maximum concentration (C_max_), average concentration (C_avg_), volume of distribution (V_z_/F), clearance (CL/F), terminal half-life (t_1/2_) of atractylodin and total AL bioactivity were calculated using non-compartmental analysis based on a linear trapezoidal rule (PKanalix version 2021R2, Antony, France: Lixoft SAS, 2021). Dose-dependent pharmacokinetic parameters were normalized with dose for evaluation of dose linearity. Data are reported as median (±ranges) values. Clinical efficacy at 4 months, including progression-free survival (PFS), overall survival (OS), and disease control rate (DCR) were assessed according to immune-related Response Evaluation Criteria in Solid tumor version [14]. The relationships between clinical responses and the pharmacokinetic parameters (AUC_0-inf_, C_max_, and C_avg_) were determined.

### 2.6 Statistical analysis

Comparisons of two independent and two dependent quantitative variables which were not normally distributed, were performed using Mann-Whitney U test and Wilcoxon Signed Rank test, respectively. Paired-t test and unpaired-t test were applied for two dependent and two independent quantitative variables, which were not normally distributed, respectively. Chi-square analysis was applied for two independent qualitative variables. Relative risk (RR) and number-need to treat (NNT) were determined following chi-square analysis. Cox proportional hazard regression model was used in the multivariate analysis to identify the predictor of disease prognosis. Akaike Information Criterion (AIC) for the four selected model parameters *(i*.*e*., AUC_0-inf_, C_max_, C_avg_, and group), -2 Log-likelihood (- 2LL) and Harrell’s C index were calculated for model performance and the goodness of fitness. Receive Operating Characteristic (ROC) analysis based on the effect of the predictor of disease prognosis was applied to determine the cut-off values of the pharmacokinetic parameters. An area Under Curve (AUC) of ≥ 0.71 indicates more predictive accuracy. Sensitivity and specificity of each cut-off value were determined. RR, sensitivity, and specificity were reported as mean ± 95% confidence interval (CI). Statistical significance level was set at α = 0.05. Cross-validation was applied to assess the internal validity of the model. All computations and analyses were performed using GraphPad Prism version 9.5 for Windows (GraphPad Software, San Diego, California, USA, www.graphpad.com). Single imputation was applied for missing data.

## 3. Results

### 3.1 Pharmacokinetic study

Data from 12 patients in group 1, 15 patients in group 2 on day 14, and 12 patients in group 2 on day 28 were available for pharmacokinetic analysis (**Figure 1**). AUC_0-inf_, AUC_0-6h_, C_max_, C_avg_, V_z_/F, CL/F, and t_1/2_ of total AL bioactivity and atractylodin in all groups are summarized in **Table 1**.

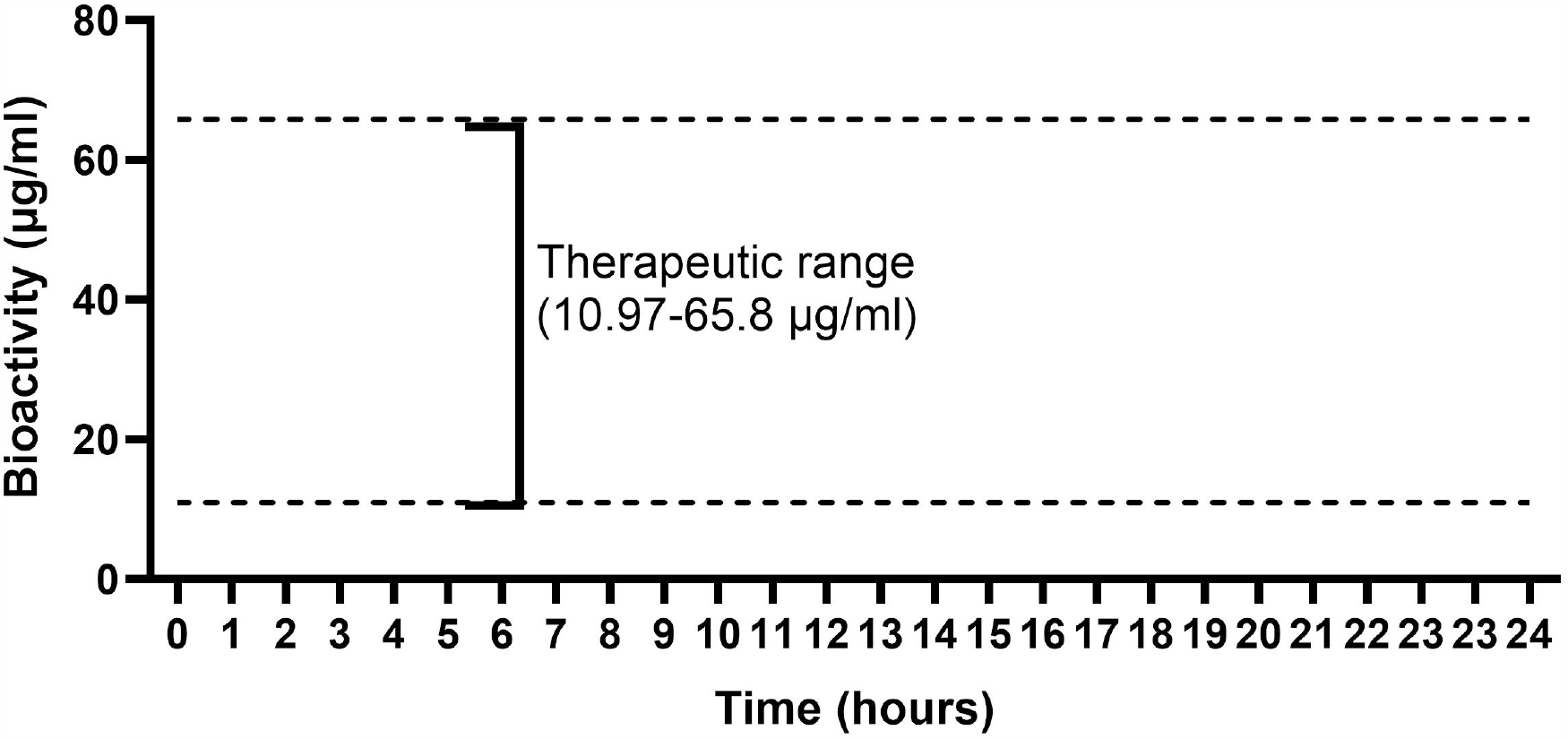

**Table 1.** Comparisons of pharmacokinetic parameters between atractylodin and total AL bioactivity. Data are presented as median (ranges).

| Pharmacokinetic parameters | Group 1 |  | Group 2 |  |  |  |
| --- | --- | --- | --- | --- | --- | --- |
|  | Day 1 |  | Day 14 |  | Day 28 |  |
|  | Atractylodin | Bioactivity | Atractylodin | Bioactivity | Atractylodin | Bioactivity |
| **AUC<sub>0-inf</sub>**<br>(ng*h/ml or *<br>µg*h/ml) | 120.07<br>(108.85-191.65) | 44.16<br>(34.11-70.46)* | 302.01<br>(221.86-408.20) | 87.73<br>(70.7-112.73)* | 443.95<br>(301.23-554.69) | 131.27<br>(95.34-170.6) |
|  | W=-76, p=0.001 |  | W=-118, p=0.0001 |  | t=7.191, df=12, p=0.0001 |  |
| **AUC<sub>0-6h</sub>**<br>(ng*h/ml or *<br>µg*h/ml) | 115.55<br>(102.42-145.88) | 38.5<br>(29.81-65.8)* | 282.59<br>(212.85-390.85) | 76.48<br>(63.46-106.28)* | 425.45<br>(262.93-499.57) | 114.57<br>(88.98-153.38)* |
|  | W=-76, p=0.001 |  | W=-118, p=0.0001 |  | t=7.289, df=12, p=0.0001 |  |
| **C<sub>max</sub>**<br>(ng/ml or *<br>µg/ml) | 44.79<br>(41.26-69.01) | 16.99<br>(15.07-20.13) * | 101.1<br>(79.77-142.67) | 34.25<br>(24.36-41.92) * | 134.99<br>(98.17-164.75) | 45.23<br>(33.47-57.37)* |
|  | W=-78, p=0.0005 |  | t=7.748, df=14, p=0.0001 |  | t=7.753, df=12, p=0.0001 |  |
| **C<sub>avg</sub>**<br>(ng/ml or *<br>µg/ml) | 19.26<br>(17.07-24.31) | 6.45 (5.70-10.93)* | 47.1 (29.23-64.85) | 12.75 (10.58-17.71) * | 70.91 (43.82-83.26) | 19.1 (14.83-25.56)* |
|  | W=-76, p=0.001 |  | W=-118, p=0.0001 |  | t=7.288, df=12, p=0.0001 |  |
| **V<sub>z</sub>/F (l)** | 573.55<br>(368.73-659.57) | 2.21<br>(1.31-2.81) | 403.93<br>(376.72-643.37) | 1.74<br>(1.25-2.19) | 399.48<br>(351.81-661.55) | 1.59<br>(1.40-1.89) |
|  | t=8.77, df=11, p=0.0001 |  | W=-120, p=0.0001 |  | t=9.258, df=12, p=0.0001 |  |
| **CL/F (l/h)** | 434.74<br>(282.54-479.96) | 1.18<br>(0.74-1.55) | 268.86<br>(198.92-366.36) | 0.92<br>(0.73-1.15) | 235.16<br>(191.69-348.16) | 0.80<br>(0.61-1.09) |
|  | t=10.74, df=11, p=0.0001 |  | t=9.614, df=14, p=0.0001 |  | t=10.05, df=12, p=0.0001 |  |
| **t<sub>1/2</sub> (h)** | 0.99<br>(0.77-1.25) | 1.24<br>(1.06-1.47) | 1.27<br>(1.04-1.47) | 1.26 (0.91-2.05) | 1.22<br>(1.15-1.62) | 1.21<br>(0.99-1.71) |
|  | W=30, p=0.2661 |  | W=26, p=0.4887 |  | t=0.4461, df=12, p=0.6635 |  |

AUC<sub>0-inf</sub>: Area Under Curve from 0h to infinity; AUC<sub>0-6h</sub>: Area Under Curve from 0h to 6h; C<sub>max</sub>: Maximum concentration; C<sub>avg</sub>: Average concentration; Df: Degree of freedom; V<sub>z</sub>/F: Volume of distribution; CL/F: Clearance; t<sub>1/2</sub>: Half-life; t: t-test; W: Wilcoxon-matched-pairs signed rank test (Sum of signed rank).

All pharmacokinetic parameters except t_1/2_ in all groups were significantly different between atractylodin and total AL bioactivity (**Table 1**). When the dose of CMC-AL was increased from 1,000 to 1,500 mg (group 1 day 1 vs. group 2 day 14), AUC_0-inf_, C_max_ and C_avg_ of atractylodin were significantly different, while t_1/2_ remained unchanged (**Table 2**). All dose-dependent and dose-independent pharmacokinetic parameters of total AL bioactivity were similar (**Table 2**). When the dose of CMC-AL was further increased from 1,500 to 2,000 mg (group 2 day 14 vs. day 28), the AUC_0-inf,_ C_max_ and C_avg_ of both atractylodin and total AL bioactivity (normalized with dose) were similar (**Table 2**).

**Table 2.** Statistical analyses for comparisons of the pharmacokinetic parameters of atractylodin and total AL bioactivity between i) group 1 on day 1 and group 2 on day 14 ii) group 2 on day 14 and group 2 on day 28.

| Pharmacokinetic parameters | Group 1 on day 1 versus Group 2 on day 14 |  | Group 2 on day 14 versus Group 2 on day 28 |  |
| --- | --- | --- | --- | --- |
|  | Atractylodin | Bioactivity | Atractylodin | Bioactivity |
| **AUC<sub>0-inf</sub>** | U=44, p=0.0246 | U=69, p=0.3229 | t=0.7684, df=26, p=0.4492 | U=87, p=0.6501 |
| **C<sub>max</sub>** | U=48, p=0.041 | U=58, p=0.1226 | t=1.193, df=26, p=0.1099 | t=0.5351, df=26, p=0.5971 |
| **C<sub>avg</sub>** | U=43, p=0.0214 | U=69, p=0.3229 | U=91, p=0.7856 | t=0.8539, df=26, p=0.401 |
| **V<sub>z</sub>/F** | t=0.4855, df=25, p=0.6316 | U=76, p=0.5164 | N/A | N/A |
| **CL/F** | U=53, p=0.0746 | t=1.027, df=25, p=0.3141 | N/A | N/A |
| **t<sub>1/2</sub>** | U=55, p=0.0903 | U=87.5, p=0.915 | N/A | N/A |

AUC<sub>0-inf</sub>: Area Under Curve from 0h to infinity; AUC<sub>0-6h</sub>: Area Under Curve from 0h to 6h; C<sub>max</sub>: Maximum concentration; C<sub>avg</sub>: Average concentration; V<sub>z</sub>/F: Volume of distribution; CL/F: Clearance; t<sub>1/2</sub>: Half-life; N/A: Not applicable

### 3.2 Identification of predictors of disease prognosis

#### 3.2.1 Progression-free survival

Multivariate cox analysis revealed no significant associations between ICC disease progression and the pharmacokinetic parameters of atractylodin or total AL bioactivity in all groups (**Table S1**). When the data from total AL bioactivity in groups 1 (day 1) and 2 (day 28) were combined, however, significant differences in the AUC_0-inf_ (t=2.78, df=20, p=0.01), C_max_ (t=3.249, df=20, p=0.004) and C_avg_ (t=2.761, df=20, p=0.01) were found between patients with the progressive and non-progressive disease. In addition, the dose-independent pharmacokinetic parameters--CL/F (U=21, p=0.016) was significantly higher in patients with progressive compared with non-progressive disease (1.18 L/h vs 0.72 L/h).

The AUC of ROC analysis revealed AUC_0-inf_ and cut-off value of 0.81 (0.6-1.0, p=0.02) and 96.71, respectively. The corresponding values for C_max_ were 0.85 (0.69-1.00, p=0.007) and 32.39, respectively, and for C_avg_ were 0.80 (0.59-1.00, p=0.02) and 14.48, respectively. The sensitivity and specificity of AUC_0-inf_ were 85.71 (60-97.46)% and 75 (40.93-95.56)%, respectively. The sensitivity [78.57 (52.41-92.43)%] and specificity [75 (40.93-95.56)%] of C_max_ and C_avg_ were equal. The RR and NNT for AUC_0-inf_ (z=2.848, p=0.004) were 3.43 (1.36-12.14) and 1.65 (1.18-8.74), respectively. Both values were similar for C_max_ (z=2.458, p=0.01) and C_avg_ (z=2.458, p=0.01) [RR=2.54 (1.18-7.15) and NNT=1.95 (1.27-25.03)].

#### 3.2.2 Inhibitory activity on tumor size progression

Multivariate cox analysis revealed no significant correlation between the increase in tumor size of ICC and the pharmacokinetic parameters of atractylodin or total AL bioactivity in all groups (**Table S2**). The dose-dependent pharmacokinetic parameters (AUC_0-inf_, C_max_, and C_avg_) of both atractylodin and total AL bioactivity were comparable in all groups (**Table S3**).

#### 3.2.3 Disease control rate (DCR)

Multivariate cox analysis revealed no significant associations between DCR and the pharmacokinetic parameters of atractylodin or total AL bioactivity in all groups (**Table S4**). When the data from group 1 (day 1) and group 2 (day 28) were combined, however, the AUC_0-inf_ (t=3.12, df=23, p=0.005), C_max_ (t=3.506, df=23, p=0.002) and C_avg_ (t=3.21, df=23, p=0.004) of total AL bioactivity were found to be a significant predictor of DCR between patients who did not respond and those who responded to treatment. Interestingly, CL/F (U=21, p=0.001) and V_z_/F (U=34, p=0.018), but not t_1/2,_ were significantly higher in responders compared with non-responders (CL/F 1.18 L/h vs 0.69 L/h and V_z_/F 2.21 L vs. 1.47 L).

The AUC of ROC analysis revealed AUC_0-inf_, C_max_ and C_avg_ of 0.83 (0.65-0.99, p=0.006), 0.84 (0.71-1.00, p=0.003) and 0.83 (0.65-0.998, p=0.006), respectively. The cut-off values for AUC_0-inf_, C_max_ and C_avg_ were 96.71, 32.39, and 14.48, respectively. The sensitivity and specificity of AUC_0-inf_ were 85.71 (60.06-97.46)% and 72.73 (43.44-90.25)%, respectively. Both parameters for C_max_ and C_avg_ were equal to AUC_0-inf_. RR and NNT for AUC_0-inf_ (z=2.961, p=0.003) were 4.0 (1.46-14.36) and 1.67 (1.22-7.05), respectively. The corresponding values for C_max_ (z=2.961, p=0.003) were close to AUC_0-inf_, and that for C_avg_ (z=2.565, p=0.01) were 2.88 (1.25-8.72) and 1.95 (1.30-15.25), respectively.

#### 3.2.3 Overall survival (OS)

Multivariate cox analysis revealed no significant associations between OS and the pharmacokinetic parameters of atractylodin or total AL bioactivity in all groups (**Table S5**). When data from group 1 (day 1) and group 2 (day 28) were combined, significant differences in OS and the AUC_0-inf_ (t=2.361, df=14, p=0.033), C_max_ (U=10, p=0.031) and C_avg_ (t=2.372, df=14, p =0.033) of total AL bioactivity were found. In addition, CL/F was significantly higher (t=2.176, df=13, p=0.04) in the deaths than in the survivors (1.22 L/h vs 0.84 L/h).

The AUC of ROC analysis revealed AUC_0-inf_, C_max_ and C_avg_ of 0.78 (0.55-1.00, p=0.06), 0.83 (0.62-1.00, p=0.03) and 0.82 (0.60-1.00, p=0.04), respectively. The cut-off values for AUC_0-inf_, C_max_ and C_avg_ were 70.46, 21.42 and 10.97, respectively. The sensitivity and specificity for AUC_0-inf_ were 83.33 (43.65-99.15)%, and 80 (49.02-96.45)%, respectively. All parameters were equal to AUC_0-inf_ for C_max_ and C_avg_. The RR and NNT for AUC_0-inf_ (z=2.472, p=0.01) were 6.43 (1.37-37.21) and 1.66 (1.16-24.15), respectively. The corresponding values for C_max_ (z=2.933, p=0.003) and C_avg_ (z=2.472, p=0.01) were 8.33 (1.79-47.58), 1.36 (1.09-6.984), 6.45 (1.36-37.21) and 1.66 (1.16-24.15), respectively.

When the data from groups 1 (day 1) and 2 (day 28) were combined, the AUC_0-inf_ and C_max_ of atractylodin were not significantly different. C_avg_, on the other hand, was considered to be a significant predictor of OS at 4 months (U=10, p=0.0312). The AUC of ROC analysis and cut-off value of atractylodin for C_avg_ were 0.83 (0.62-1.00, p=0.03) and 22.10, respectively. The sensitivity and specificity were 83.33 (43.65-99.15)%, and 80 (49.02-96.45)% respectively. RR and NNT (z=2.472, p=0.01) were 6.45 (1.36-37.21) and 1.66 (1.16-24.15), respectively. CL/F was significantly lower (t=2.257, df=14, p=0.04) in the survivors than in the deaths (302.45 vs. 433 L/h). AUC_0-inf_, C_max_ and C_avg_ of atractylodin or total AL bioactivity in other groups were comparable.

### 3.3 Cross-validation

Since there were no significant associations between clinical responses (PFS, DCR, and OS) and the pharmacokinetics of total AL bioactivity (AUC_0-inf_, C_max_, and C_avg_) in group 2 patients on day 14 (internal validation), cross-validation was applied for internal validity of the model to confirm that the prognostic factors can be applied to other scenarios.

#### 3.3.1 Progressive disease

The sensitivity and specificity for AUC_0-inf_ were 66.67 (30-94.08)%, and 83.33 (43.65-99.15)%, respectively. The corresponding values for C_max_ were 50 (18.76-81.24)%, and 83.33 (43.65-99.15)%, respectively. All values for C_avg_ were equal to C_max_.

#### 3.3.2 DCR

The sensitivity and specificity for AUC_0-inf_ were 66.67 (30-94.08)% and 85.71 (48.69-99.27)%, respectively. The corresponding values for C_max_ were 50 (18.76-81.24)% and 85.71 (48.69-99.27)%, respectively. These values for C_avg_ were equal to C_max._

#### 3.3.3 OS

There were 3 patients who survived at 4 months of treatment. The sensitivity and specificity for AUC_0-inf_ were 100 (56.55-100)% and 0%, respectively. The corresponding values for C_max_ were 100 (56.55-100)% and 33.33 (1.7-88.15)%, respectively. These values for C_avg_ were equal to C_max_.

## 4. Discussion

The prognosis models for the PFS, DCR and OS associated with CMC-AL therapy in patients with advanced-stage ICC were built with cross-validation. The models were presented as independent predictors of disease prognosis with cut-off values. The therapeutic ranges of atractylodin and total AL bioactivity were also determined. The study was the first that applied total bioactivity, instead of a single active constituent, as a marker of pharmacokinetic profiles following treatment with herbal medicine. This approach is convincing as herbal medicine may consist of a mixture of various active constituents, some of which are unidentified.

### 4.1 Pharmacokinetic study

The pharmacokinetic parameters of atractylodin and total AL bioactivity on day 28 after continuous dosing, indicating the accumulation of AL was unlikely. This could be due to the short half-life of atractylodin and total AL bioactivity. More frequent dosing, *i*.*e*., twice/thrice/fourth daily dosage regimens, would be a favourable choice to maintain plasma/tissue concentrations of AL at the target sites of AL action. Further phase 2B clinical trial is required to confirm the optimal dose regimens of CMC-AL in patients with advanced-stage ICC to avoid an excessive number of capsules administered for each dose. The pharmacokinetics of atractylodin following the once-daily dose of 1,000 mg of CMC-AL observed in advanced-stage ICC patients was in agreement with the previous reports in the phase 1 study in healthy volunteers [12], suggesting no influence of ICC disease on the pharmacokinetics of atractylodin. Therefore, the determined dosage regimen of CMC-AL in patients with advanced-stage ICC can be applied to all stages of the patients. Total AL bioactivity showed dose linearity over the dose range of 1,000 to 2,000 mg. On the other hand, atractylodin showed dose non-linearity over the range 1,000 to 2,000 mg. All the dose-independent pharmacokinetics of atractylodin and total AL bioactivity were similar over this dose range.

### 4.2 Predictors of disease prognosis

#### 4.2.1 Progression-free survival (PFS)

Based on the ROC analysis for the PFS at 4 months, all of the dose-dependent pharmacokinetic parameters were promising predictors of disease progression (AUC≥0.71), with high predictive accuracy. AUC_0-inf_ provided the best sensitivity and specificity (AUC=0.81). Although the AUC of C_max_ was highest, the sensitivity and specificity of C_max_ were lower than AUC_0-inf_. The AUC_0-inf_ was, therefore, the preferable choice as a predictor of PFS at 4 months (**Figure 2**). The suggested AUC_0-inf_ (cut-off) was > 96.71 µg*h/ml to improve PFS in advanced-stage ICC patients with CMC-AL treatment. For cross-validation, the sensitivity of AUC_0-inf_ was slightly decreased, while the specificity was increased, indicating high predictive accuracy. It was obvious that the sensitivity of C_max_ and C_avg_ was decreased to 50%, while the specificity was increased to 83.33%. C_max_ and C_avg_ could be optional predictors of ICC prognosis. In the previous reports, no association between C_max_ of total AL bioactivity and the incidence of adverse reaction were found [11]. The therapeutic windows of total AL bioactivity for PFS range from 14.48 (cut-off value for C_avg_) to 65.8 µg/ml. It was noted for the higher CL/F of total AL bioactivity in patients with disease progression than those without disease progression. Pharmacogenetics of drug-metabolizing enzymes and/or protein transporters may be involved. Further study focusing on genetic polymorphisms of drug-metabolizing enzymes and/or protein transporters would assist in determining the predefined criteria for CMC-AL treatment in ICC. Such high CL/F in patients with disease progression resulted in insufficient AUC_0-inf_, C_max_ and C_avg_ that could lead to treatment failure due to suboptimal drug concentrations in plasma and target tissues. Besides genetic polymorphisms, high CL/F in patients with progression disease may be due to induction of drug metabolizing enzymes during the on-going phase of disease progression. Significant influence of renal function is unlikely since there was no significant difference in estimated glomerular filtration rate (eGFR) between those with and without disease progression [13].The results from this study suggest that AUC_0-inf_ is the best surrogate predictor of disease prognosis for disease progression in patients with advanced-stage ICC. C_max_ and C_avg_ could be used as surrogates for treatment optimization. Patients with AUC_0-inf_ of < 96.71 µg*h/ml were associated with an increased risk of disease progression of up to 12.14-fold (RR: 3.43, 95%CI: 1.36-12.14) compared with those with AUC_0-inf_ of > 96.71 µg*h/ml. The risk of disease progression was also increased in patients with C_max_ of < 32.39 µg/ml and C_avg_ of < 14. 48 µg/ml, although the contribution was less pronounced when compared with AUC_0-inf_ (RR: 2.54, 95%CI: 1.18-1.25). The NNT values (1.65, 1.95, 1.95 for AUC_0-inf_, C_max_ and C_avg_, respectively) indicate a large effect size difference between the patients who have these pharmacokinetic parameters above or below the cut-off levels. This suggests that progression-free disease is expected to be observed in every two patients who received treatment with CMC-AL who have AUC_0-inf_, C_max_, and C_avg_ above the cut-off values or there’s a 50 percent chance that those with AUC_0-inf_, C_max_, and C_avg_ above the cut-off point will prevent the disease progression. CMC-AL showed potential anti-CCA activity in reducing the risk of disease progression in advanced-stage ICC patients. First-line therapy with conventional drugs gemcitabine/cisplatin combination was reported to reduce the risk of disease progression by up to 1.96-fold (1.65, 95%CI: 1.35-1.96) compared with gemcitabine alone in patients with advanced-stage CCA. Targeted therapy with ivosidenib (an isocitrate dehydrogenase-1 inhibitor) on the other hand, reduced the risk of disease progression by 4-fold (2.7, 95%CI: 1.85-4) compared with placebo [15]. A study of regorafenib in patients who failed gemcitabine/platinum-based therapy showed that the risk of disease progression was reduced by up to 3.44-fold (2.04, 95%CI: 1.23-3.44) compared with placebo [16]. In addition to targeted therapy, a recent study of immunotherapy reported that patients treated with durvalumab in combination with gemcitabine in patients with advanced-stage CCA had a significant decrease in the risk of disease progression by up to 1.59-fold (1.33, 95%CI: 1.12-1.59) compared with placebo [17].

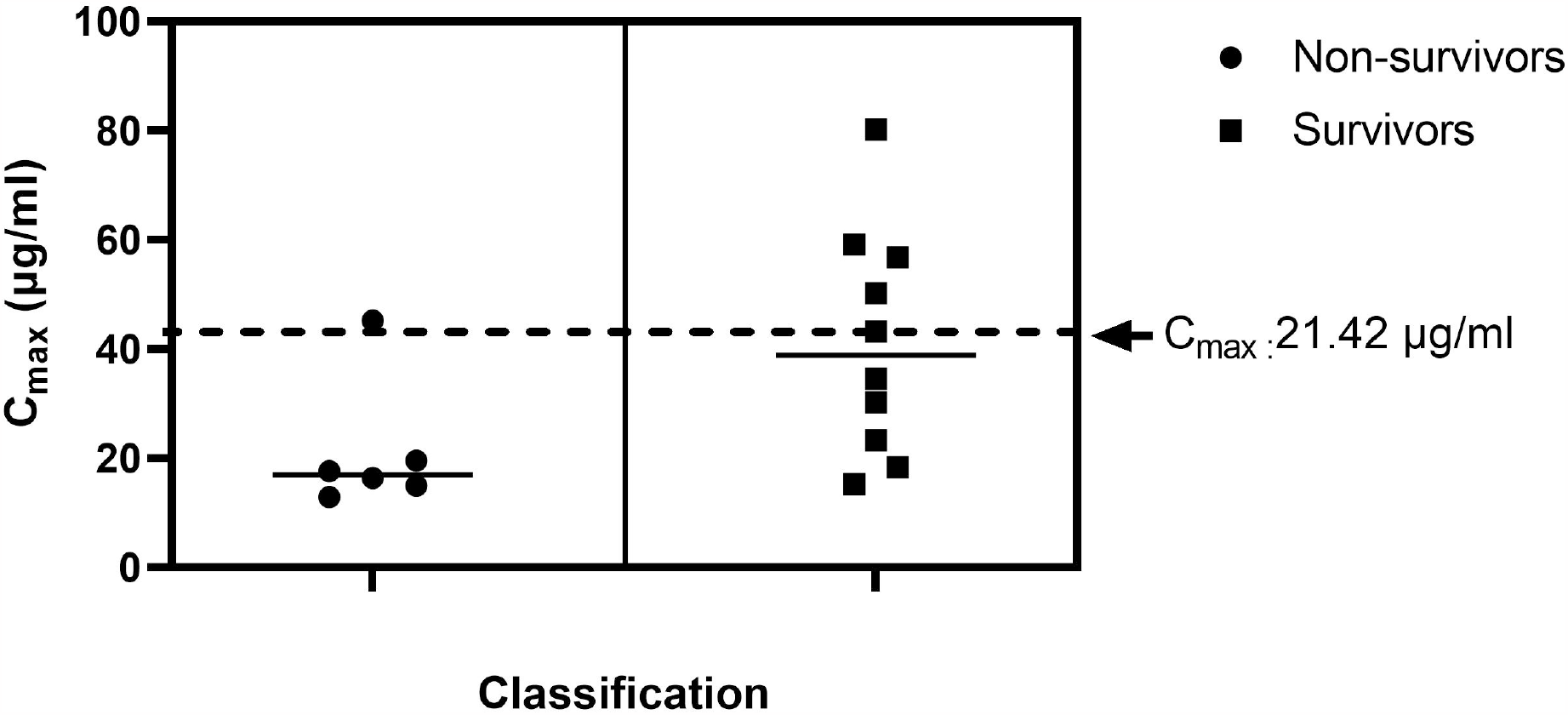

#### 4.2.2 DCR

Similarly to PFS, AUC_0-inf_ was the best predictor of disease prognosis associated with DCR, although sensitivity and specificity were equal to C_max_ and C_avg_. Based on ROC analysis, the AUC of AUC_0-inf_ (0.81) was lower than C_max_ (0.84), but was still considered highly accurate (**Figure 3**). However, the sensitivity (66.67%) and specificity (85.71%) of the validated AUC_0-inf_ was the highest.

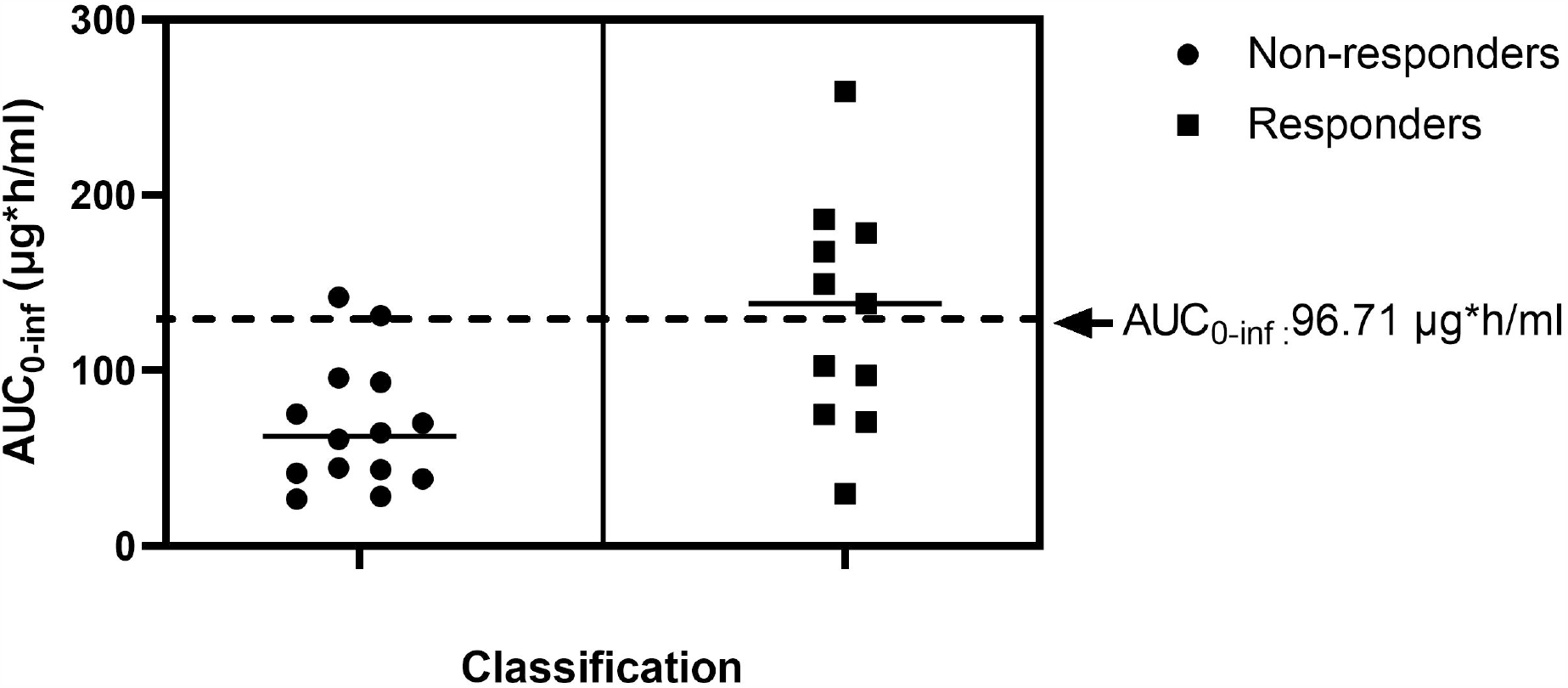

The sensitivity of C_max_ and C_avg_ was on the other hand, was dramatically decreased to 50%. Furthermore, the RR for AUC_0-inf_ was up to 14-fold (4, 95%CI: 1.46-14.36) to increase DCR, while the RR for C_avg_ was up to 8.72-fold (2.88, 95%CI: 1.25-8.72). Hence, the increased in DCR for AUC_0-inf_ was 1.64-fold higher than C_avg_. The NNT of AUC_0-inf_ was lower than C_avg_, (1.67 vs. 1.95). The NNT for the success of treatment based on the prognostic predictor of AUC_0-inf_ was also only 2, which indicates a large effect size difference between the two groups. Although C_avg_ may not be a favorable predictor of DCR, it could be used to identify therapeutic window of total AL bioactivity (14.48 to 65.8 µg/ml).

Similarly to PFS, CL/F and V_z_/F were significantly different between the responders and non-responders based on the DCR criteria. Apart from the contribution of pharmacogenetic factors that resulted in accelerated CL/F, physiologic factors such as the presence of ascites or obesity may have resulted in the expansion of the volume of distribution and thus, suboptimal systemic drug exposure. Patients with ascites or obesity may therefore, require a high dose regimen of CMC-AL.

#### 4.2.3 OS

In contrast to PFS and DCR, the most promising predictor of prognosis for OS was C_max_ (**Figure 5**). This was supported by the ROC analysis (AUC >0.71), sensitivity, and specificity. In addition, the sensitivity and specificity obtained from the cross-validation of C_max_ was higher than AUC_0-inf_. Alternatively, C_avg_ could be applied. With the highest RR, patients with C_max_ of > 21.42 µg/ml had an increased OS by up to 47.58-fold (8.33, 95%CI: 1.79-47.58) compared with those with C_max_ of < 21.42 µg/ml. Patients with AUC_0-inf_ of < 70.86 µg*h/ml, on the other hand, had an increased OS by up to 37.21 (6.43, 95%CI: 1.37-37.21) compared with those with AUC_0-inf_ of <70.86 µg*h/ml. In addition, the NNT for C_max_ was lower than AUC_0-inf_ (1.36 vs. 1.66). In case where the measurement of total AL bioactivity was not available, C_avg_ of atractylodin would be an optional choice as it provided comparable AUC to C_max_ of total AL bioactivity, although the RR and NNT values were relatively lower (only up to 37.25 and 1.65, respectively). Interestingly, the cut-off value for OS was lower than PFS and DCR, indicating the requirement of lower drug concentrations to obtain desirable therapeutic outcome. Thus, the therapeutic window of total AL bioactivity for OS ranged from 10.97 (cut-off value of C_avg_) to 65.8 µg/ml (**Figure 6**). CL/F was also higher in the non-survivors (deaths) compared with survivors, indicating the significant contribution of drug clearance in the non-survivors. In-depth analysis of the association between drug clearance in non-survivors is necessary to enhance the clinical efficacy of CMC-AL therapy in patients with advanced-stage ICC.

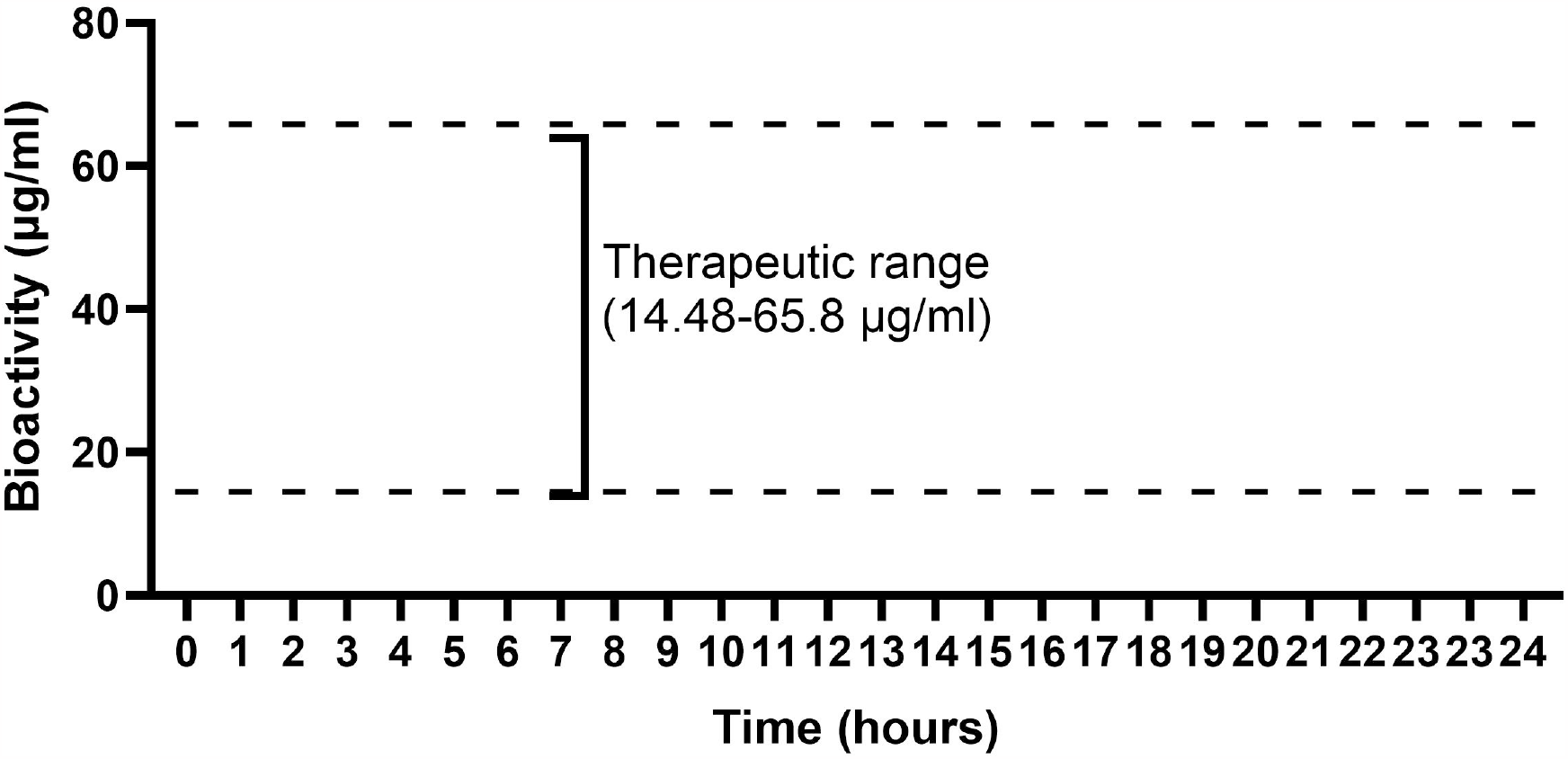

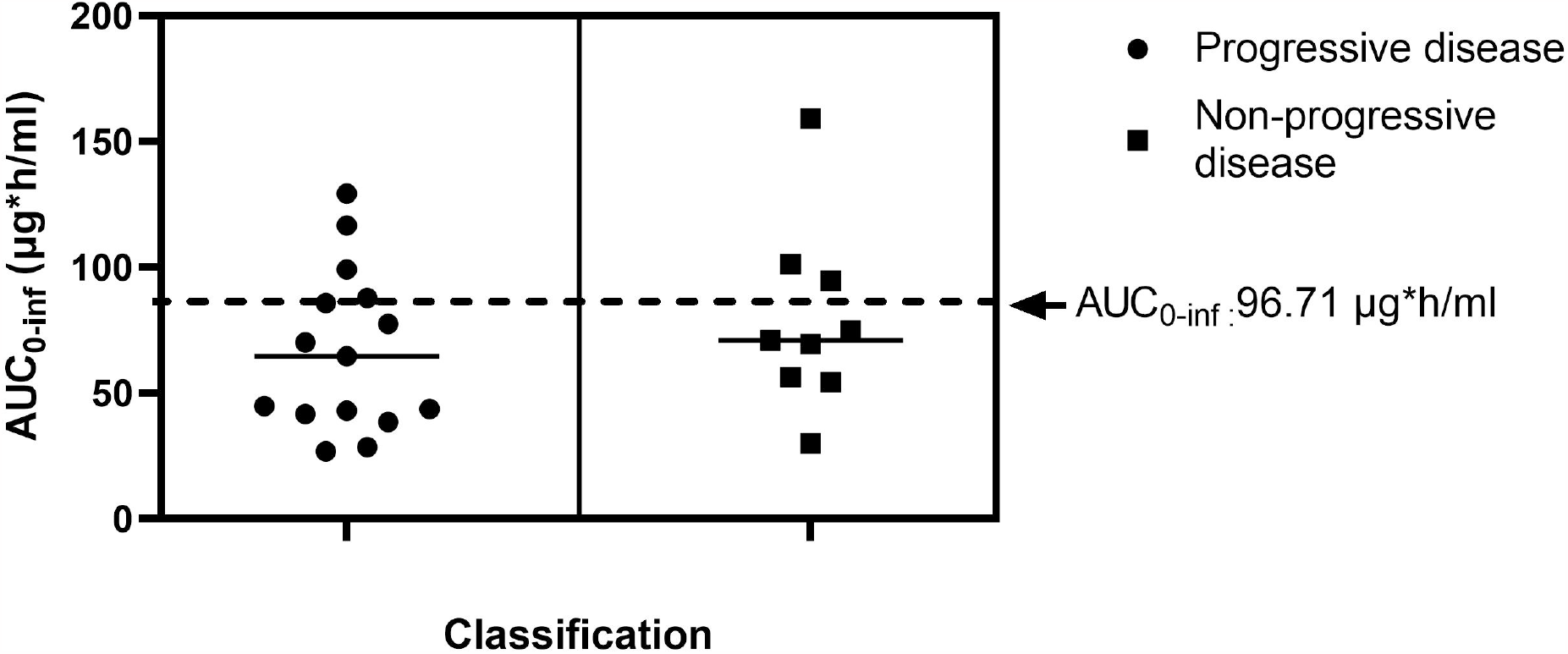

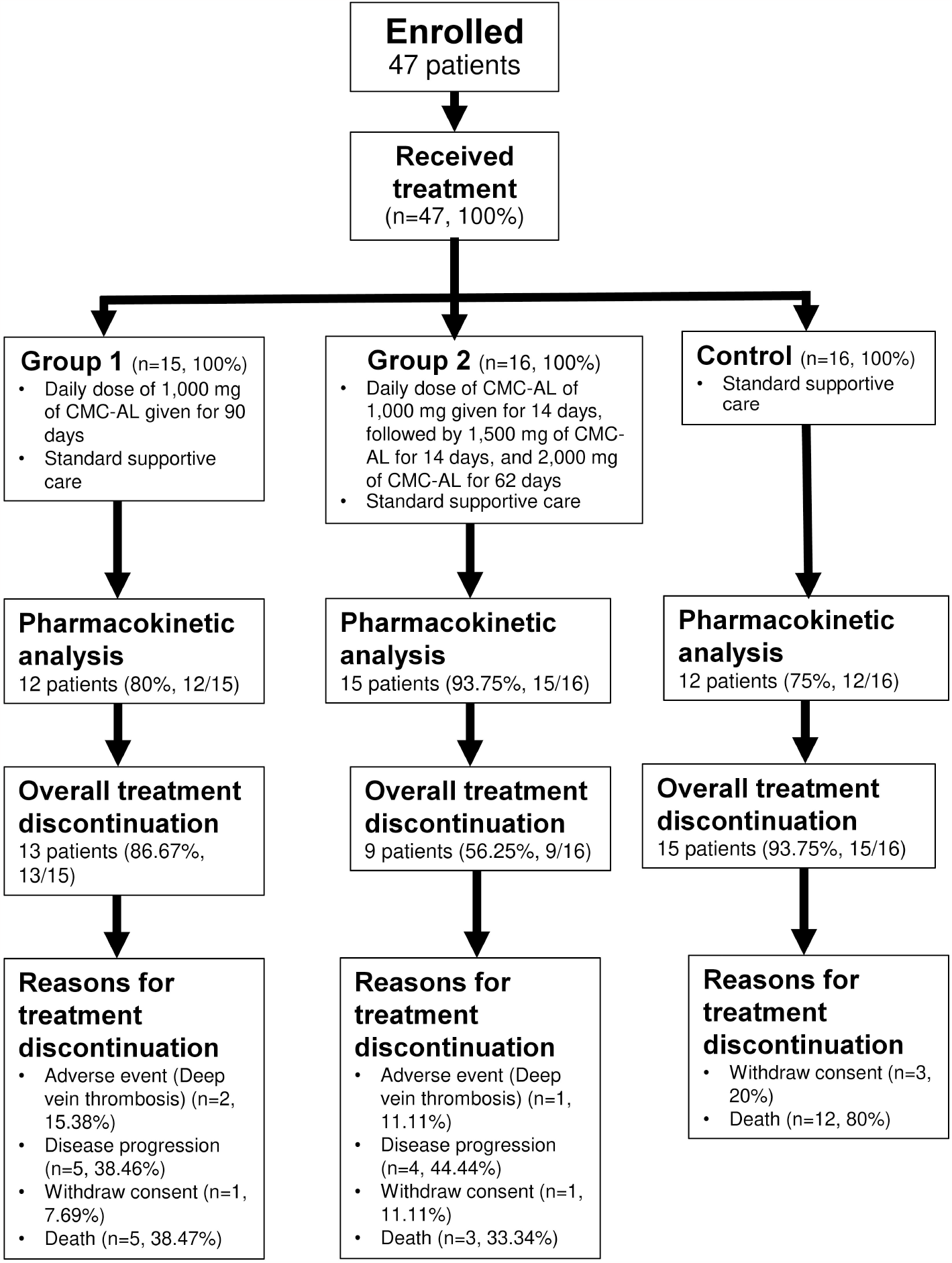
Capsule formulation of the standardized AL extract (CMC-AL)

It was evident for the potential of CMC-AL to increase the OS in ICC patients with advanced-stage ICC. Gemcitabine/cisplatin combination therapy was shown to prevent the risk if disease progression by up to 3-fold (1.59; 95%CI: 1.30-3.0) in patients with advanced-stage CCA [18]. The second-line therapy --capecitabine on the other had did not reduce the risk of death (hazard ratio (HR): 0.81, 95%CI: 0.63-1.04) in patients with advanced-stage CCA [19]. The FOLFOX regimen containing folinic acid, fluorouracil, and oxaliplatin, on the other hand, was reported to reduce the risk of death by up to 2-fold (1.44, 95%CI: 1.03-2) compared with control [20]. Patients with immunotherapy--durvalumab in combination with gemcitabine was reported to increase the survival rate of patients with advanced-stage CCA by up to 1.5-fold (1.25, 95%CI: 1.03-1.5) [17].

In conclusion, the predictors of ICC disease prognosis (*i*.*e*., AUC_0-inf_, C_max,_ and C_avg_) were successfully established with different cut-off values to improve PFS, DCR and OS in patients with advanced-stage ICC who were treated with CMC-AL. In addition, the therapeutic ranges of total AL bioactivity were determined. Notably, the study highlights the importance of the measurement of pharmacokinetic parameters of total AL bioactivity since these parameters were clinically correlated with clinical outcomes rather than a single active constituent -- atractylodin. This approach should be applied to other herbal medicines to determine the pharmacokinetic-pharmacodynamic relationship and therapeutic window. The limitation of the study is the low number of participants recruited in each group, as well as the lack of external validation. Therefore, the prediction of ICC disease prognosis using the proposed surrogates may not be applied to all scenarios. In addition, the small sample size resulted in lower sensitivity and specificity in the cross-validation, which may have had a direct effect on model accuracy. Further study for an external validation with larger sample size is needed to confirm the model accuracy and applicability.

## Data Availability

The authors confirm that the data supporting the findings of this study are available within the article and/or its Supplementary Materials and methods. Any additional data are available from the corresponding author K.N. upon reasonable request.

## Additional information

Supplementary Information the online version contains supplementary material available at.

## Credit authorship contribution statement

T.S. designed the experiment. T.S., J.K., A.C., and T.P. performed the experiment. T.S. and J.K. analyzed the data. T.S. wrote the original draft paper. T.S. provided the program for analysis. K.N. supervised all study. K.N. and J.K. did a critical revision. All authors contributed to the discussion of results and manuscript corrections.

