## Supplementary information for "Prognostic factors of clinical responses in patients with advanced-stage intrahepatic cholangiocarcinoma following *Atractylodes lancea* administration: A phase 2A clinical trial"

**Table S1.** Multivariate cox analysis between progressive disease of intrahepatic cholangiocarcinoma and pharmacokinetic parameters of atractylodin or total AL bioactivity

| **Parameters** | **GR1_D1_GR2_D14** | | **GR1_D1_GR2_D28** | |
| --- | --- | --- | --- | --- |
|  | **ATD** | **Bioactivity** | **ATD** | **Bioactivity** |
| **Group** | Z=0.7952, p=0.4265 | Z=0.552, p=0.581 | Z=0.5628, p=0.5736 | Z=1.248, p=0.212 |
| **AUC_0-inf_** | Z=1.058, p=0.2899 | Z=1.181, p=0.2375 | Z=0.8062, p=0.4202 | Z=1.164, p=0.2443 |
| **C_max_** | Z=0.4989, p=0.6179 | Z=0.4458, p=0.9644 | Z=0.3465, p=0.7289 | Z=1.502, p=0.133 |
| **C_avg_** | Z=1.274, p=0.2025 | Z=1.31, p=0.1904 | Z=0.8639, p=0.3877 | Z=0.8439, p=0.3987 |
| **AIC** | 31.75 | 31.75 | 27.23 | 30.72 |
| **-2LL** | 23.75 | 23.75 | 19.13 | 22.72 |
| **C-index** | 0.71 (0.37-1.04) | 0.71 (0.52-0.89) | 0.62 (0.23-1.00) | 0.73 (0.49-0.96) |
| **Hypothesis (Score test)** | 3.961, p=0.4113 | 3.264, p=0.5147 | 1.932, p=0.7482 | 3.158, p=0.5317 |
| **Is the model correct?** | Incorrect | Incorrect | Incorrect | Incorrect |

AUC_0-inf_: Area Under Curve from 0h to infinity; ATD: Atractylodin; AIC: Akaike Information Criterion (AIC) for four selected model parameters,-2LL: -2 Log-likelihood (-2LL); C-index: Harrell’s C-index; C_max_: Maximum concentration; C_avg_: Average concentration;

**Table S2** Multivariate cox analysis between increased in tumor size of intrahepatic cholangiocarcinoma and pharmacokinetic parameters of atractylodin or total AL bioactivity

| **Parameters** | **GR1_D1_GR2_D14** | | **GR1_D1_GR2_D28** | |
| --- | --- | --- | --- | --- |
|  | **ATD** | **Bioactivity** | **ATD** | **Bioactivity** |
| **Group** | Z=1.772, p=0.0765 | Z=0.7626, p=0.4457 | Z=1.569, p=0.1165 | Z=0.3299, p=0.7415 |
| **AUC_0-inf_** | Z=1.667, p=0.0956 | Z=1.396, p=0.1626 | Z=1.221, p=0.223 | Z=0.5517, p=0.5812 |
| **C_max_** | Z=1.537, p=0.1244 | Z=1.548, p=0.1216 | Z=0.7941, p=0.4272 | Z=1.346, p=0.1783 |
| **C_avg_** | Z=1.375, p=0.169 | Z=1.061, p=0.2889 | Z=1.447, p=0.148 | Z=0.2399, p=0.8104 |
| **AIC** | 91.43 | 30.74 | 87.86 | 88.41 |
| **-2LL** | 83.43 | 22.74 | 79.86 | 80.41 |
| **C-index** | 0.68 (0.55-0.80) | 0.66 (0.39-0.93) | 0.65 (0.51-0.79) | 0.51 (0.35-0.67) |
| **Hypothesis (Score test)** | 6.705, p=0.1523 | 3.951, p=0.4127 | 5.136, p=0.2736 | 3.711, p=0.4466 |
| **Is the model correct?** | Incorrect | Incorrect | Incorrect | Incorrect |

AUC_0-inf_: Area Under Curve from 0h to infinity; ATD: Atractylodin; AIC: Akaike Information Criterion (AIC) for four selected model parameters,-2LL: -2 Log-likelihood (-2LL); C-index: Harrell’s C-index; C_max_: Maximum concentration; C_avg_: Average concentration;

**Table S3.** Comparisons of dose-dependent pharmacokinetic parameters of atractylodin and total bioactivity between increased in tumor size (tumor progression) and non-increased in tumor size (decreased in tumor size or stable in tumor size)

| **Parameters** | **GR1_D1_GR2_D14** | | **GR1_D1_GR2_D28** | |
| --- | --- | --- | --- | --- |
|  | **ATD** | **BIOACTIVITY** | **ATD** | **BIOACTIVITY** |
| **AUC_0-inf_** | U=66, p=0.621 | U=74, p=0.9379 | U=48, p=0.389 | t=0.6678, df=23, p=0.5109 |
| **C_max_** | U=71, p=0.815 | U=70, p=0.7662 | U=56, p=0.7007 | U=40.5, p=0.31 |
| **C_avg_** | U=63, p=0.5147 | U=73, p=0.8966 | U=47, p=0.3567 | t=0.6846, df=23, p=0.5004 |

AUC_0-inf_: Area Under Curve from 0h to infinity; ATD: Atractylodin; C_max_: Maximum concentration; C_avg_: Average concentration; U: Mann Whitney U test

**Table S4** Multivariate cox analysis between increased in disease control rate (DCR) cholangiocarcinoma and pharmacokinetic parameters of atractylodin or total AL bioactivity

| **Parameters** | **GR1_D1_GR2_D14** | | **GR1_D1_GR2_D28** | |
| --- | --- | --- | --- | --- |
|  | **ATD** | **Bioactivity** | **ATD** | **Bioactivity** |
| **Group** | Z=0.4567, p=0.6479 | Z=0.1825, p=0.8552 | Z=0.067, p=0.9462 | Z=1.716, p=0.0863 |
| **AUC_0-inf_** | Z=2.254, p=0.0242 | Z=0.6784, p=0.4975 | Z=1.112, p=0.2662 | Z=0.1341, p=0.8933 |
| **C_max_** | Z=1.883, p=0.0597 | Z=0.8206, p=0.4119 | Z=0.3415, p=0.7327 | Z=0.7042, p=0.4813 |
| **C_avg_** | Z=2.051, p=0.0403 | Z=0.2371, p=0.8126 | Z=0.7375, p=0.4608 | Z=0.6275, p=0.5303 |
| **AIC** | 38.08 | 42.67 | 37.66 | 38.36 |
| **-2LL** | 30.08 | 34.67 | 29.66 | 30.36 |
| **C-index** | 0.82 (0.64-1.00) | 0.64 (0.44-0.87) | 0.61 (0.30-0.91) | 0.72 (0.50-0.94) |
| **Hypothesis (Score test)** | 4.021, p=0.4031 | 7.903, p=0.0952 | 2.235, p=0.6927 | 8.861, p=0.0647 |
| **Is the model correct?** | Incorrect | Incorrect | Incorrect | Incorrect |

AUC_0-inf_: Area Under Curve from 0h to infinity; ATD: Atractylodin; AIC: Akaike Information Criterion (AIC) for four selected model parameters,-2LL: -2 Log-likelihood (-2LL); C-index: Harrell’s C-index; C_max_: Maximum concentration; C_avg_: Average concentration;

**Table S5.** Multivariate cox analysis between increased in overall survival of intrahepatic cholangiocarcinoma and pharmacokinetic parameters of atractylodin or total AL bioactivity

| **Parameters** | **GR1_D1_GR2_D14** | | **GR1_D1_GR2_D28** | |
| --- | --- | --- | --- | --- |
|  | **ATD** | **Bioactivity** | **ATD** | **Bioactivity** |
| **Group** | Z=0.3899, p=0.6966 | Z=0.5235, p=0.6006 | Z=0.1888, p=0.8503 | Z=0.1821, p=0.8555 |
| **AUC_0-inf_** | Z=0.8985, p=0.3689 | Z=0.7842, p=0.4329 | Z=0.7855, p=0.4322 | Z=0.4208, p=0.6739 |
| **C_max_** | Z=0.4346, p=0.6639 | Z=0.3944, p=0.6933 | Z=0.6751, p=0.4996 | Z=0.1529, p=0.8785 |
| **C_avg_** | Z=0.9643, p=0.3349 | Z=0.8169, p=0.414 | Z=0.7232, p=0.4696 | Z=0.1631, p=0.8704 |
| **AIC** | 44.29 | 45.77 | 33.94 | 33.72 |
| **-2LL** | 36.29 | 37.77 | 25.94 | 25.72 |
| **C-index** | 0.68 (0.49-0.86) | 0.69 (0.47-0.90) | 0.75 (0.55-0.94) | 0.76 (0.53-0.99) |
| **Hypothesis (Score test)** | 4.649, p=0.3253 | 3.241, p=0.5183 | 4.502, p=0.3423 | 4.287, p=0.3685 |
| **Is the model correct?** | Incorrect | Incorrect | Incorrect | Incorrect |

AUC_0-inf_: Area Under Curve from 0h to infinity; ATD: Atractylodin; AIC: Akaike Information Criterion (AIC) for four selected model parameters,-2LL: -2 Log-likelihood (-2LL); C-index: Harrell’s C-index; C_max_: Maximum concentration; C_avg_: Average concentration;
